# Digital Twin Workflow for Continuous Glucose Forecasting: Real-Time Data Assimilation Using Reinforcement Learning

**DOI:** 10.64898/2026.10.23.26366163

**Authors:** Xiang Cao, Qi Wang

## Abstract

Digital twins for physiological forecasting require robust mechanisms for continuously assimilating streaming data and adapting to patient-specific dynamics. Threshold-triggered model updates can restore predictive performance after substantial drift, but their reactive nature limits continuous adaptation. We propose a proactive digital twin framework that couples a pretrained PatchTST foundation model with an Actor–Critic reinforcement learning (RL) agent for residual correction. Using a 30-minute look-back window and recent residual history, the agent generates bounded corrections (±0.05) to the first-step baseline forecast, with corrected predictions recursively propagated over a 15- step horizon. We evaluate both an RL-only workflow and a hybrid workflow combining RL correction with threshold-triggered fine-tuning using minute-by-minute continuous glucose monitoring data from 10 critically ill patients with sepsis. Across all patients, the pretrained RL agent improves forecasting accuracy without patient-specific retraining. Both workflows reduce trajectory-averaged mean absolute percentage error, ensemble relative error, and maximum errors compared with the uncorrected baseline and previously developed reactive digital twin models. These findings support residual reinforcement learning as a lightweight mechanism for continuous, real-time data assimilation that preserves the foundation model’s global predictive prior while enabling patient-specific forecast adaptation, with potential applications in critical care decision support.

## 1 Introduction

Sepsis presents a major challenge in intensive care because of acute organ dysfunction, rapid physiological deterioration, and substantial inter-patient variability in disease progression and treatment response. Glucose dysregulation is a central metabolic manifestation of sepsis and critical illness, arising from stress-hormone activation, inflammatory signaling, insulin resistance, altered hepatic glucose production, and changes in nutritional and pharmacological support. Both hyperglycemia and hypoglycemia are associated with adverse outcomes in critically ill patients. Moreover, evidence that overly intensive glucose control can increase harm highlights the need for individualized forecasting systems that can support careful monitoring of evolving glucose dynamics [5, 8].

Continuous glucose monitoring (CGM) offers new opportunities for real-time forecasting and bedside decision support in hospital and intensive care unit (ICU) settings. Compared with intermittent blood sampling, CGM provides high-frequency measurements that reveal short-term variability, directional trends, and glucose excursions that might otherwise go undetected. These capabilities are particularly valuable for critically ill patients, whose glucose levels can change rapidly in response to infection, organ dysfunction, nutrition, and insulin administration [4,7]. However, access to high-resolution observations alone is insufficient for reliable forecasting. A predictive system must also assimilate these observations and remain synchronized with the changing dynamics of each patient.

Digital twins provide a framework for meeting this requirement. In healthcare, a digital twin is a patient-specific computational representation that is continuously updated with incoming data to predict future states under changing clinical conditions. Recent reviews identify personalization, dynamic data connection, and predictive capability as defining features of digital twins intended to support clinical decisions over time [3, 6, 10]. These features are especially relevant in the ICU, where individual trajectories may rapidly diverge from population-level patterns. A digital twin for glucose forecasting must therefore combine a predictive model with an effective mechanism for assimilating streaming patient data.

In our previous work, we developed a digital twin workflow for patients with sepsis using a pretrained PatchTST foundation model and threshold-triggered transfer learning. The model was fine-tuned on patient-specific data when the forecasting error exceeded a prescribed threshold. This approach demonstrated the feasibility of patient-specific model adaptation, but its updating mechanism remained reactive: recalibration occurred only after a sufficiently large discrepancy had developed. Although retraining can restore predictive performance after drift, threshold-triggered parameter updates do not themselves provide continuous forecast correction. This limitation motivates an adaptation mechanism that responds to new observations at each prediction cycle while retaining the temporal structure learned by the pretrained model. Reinforcement learning (RL) offers a way to learn such a correction mechanism. An RL agent can use recent glucose observations and prediction errors to adjust the forecast through a continuous action, without repeatedly retraining the forecasting backbone. Actor–Critic methods are well suited to this formulation because they support continuous actions and optimize both immediate and longer-term rewards [9]. More broadly, RL has shown promise for sequential decision-making under uncertainty in healthcare, where an agent interacts with an evolving system rather than making an isolated prediction [11]. Applied to digital twin forecasting, this suggests complementary roles for the two components: the foundation model captures the temporal structure of glucose dynamics, and the RL policy provides patient-specific residual corrections based on recent observations and model errors.

In this paper, we develop two related digital twin workflows for continuous glucose forecasting. The first couples a frozen PatchTST foundation model with an Actor–Critic agent. At each prediction time, the agent observes a rolling glucose window and the recent residual history, then applies a bounded correction to the first-step baseline forecast. The corrected value initializes a recursive multi-step rollout generated by the foundation model. Clipping the correction to a prescribed range and penalizing its magnitude in the reward function constrain the adjustment. This design enables continuous forecast adaptation while keeping both the pretrained backbone and the deployed RL policy fixed. The second workflow combines this residual-correction mechanism with threshold-triggered transfer learning. Its inner loop adjusts forecasts at each time step, while its outer loop monitors the ensemble forecasting error and fine-tunes the backbone when an update is triggered.

We evaluate these workflows using CGM data from critically ill patients and compare them with the uncorrected PatchTST model and the previous transfer-learning-based digital twin. The evaluation examines the contribution of residual correction, the additional effect of backbone updating, and the behavior of an alternative update criterion. The pretrained RL agent improves one-step forecasting accuracy for all 10 evaluation patients without further patient-specific retraining. The hybrid workflow reduces the mean ensemble relative error in nine patients and the maximum error in all 10, although backbone fine-tuning does not provide additional gains over RL-only correction in the patients for whom it is triggered. These findings clarify the benefits of continuous residual assimilation and the challenges of coordinating a fixed correction policy with an updated predictive backbone.

The remainder of the paper is organized as follows. Section 2 describes the datasets, reviews the baseline digital twin workflow, and presents the RL-based residual-correction and hybrid methods. Section 3 reports the numerical results and comparisons. Sections 4 and 5 discuss the findings and present the conclusions, respectively.

## 2 Methods

### 2.1 Datasets for building the foundation model and digital twins

The data used to build the foundation model (or template) were obtained from a middle aged woman with sepsis and diabetes who was admitted to Sichuan Provincial People’s Hospital with sudden dysarthria, convulsions, and loss of consciousness [2]. Following enrollment, she was fitted with a continuous glucose monitoring (CGM) device (FreeStyle Libre, Abbott Diabetes Care Ltd., UK). Glucose data from the preceding 24 hours were downloaded each morning at 8:00 AM. Upon discharge from the emergency intensive care unit (EICU), the device was removed and stored for centralized data collection. The dataset comprises minute-by-minute glucose measurements collected from a 4-day period three years ago, yielding 19,621 data points. Using the foundation model trained on this source patient’s data, we developed digital twins to monitor and forecast glucose levels in individual patients with sepsis. We evaluated the workflow on 10 additional patients with sepsis. Their clinical data included demographics, admission characteristics, and clinical scores (APACHE II, SOFA, and NRS-2002) obtained from the Emergency Medical Record System. Laboratory and biochemical data were retrieved from the Laboratory Information System, and nutritional and insulin administration records were obtained from the Hospital Information System. Continuous physiological variables, including ventilation, infusion, and ECG data, were captured in real time through the EICU automated monitoring platform. The CGM system provided minute-by-minute glucose readings throughout the ICU stay; these data were downloaded daily and archived upon discharge. Because insulin data had a limited impact on glucose forecasting in our previous study [2], the digital twins developed here use only CGM glucose data.

The collection and use of these data formed part of an observational study. The study protocol was approved by the Medical Ethics Committee of Sichuan Provincial People’s Hospital and Sichuan Academy of Medical Sciences (approval number: LunShen(Yan)-2026-250) in March 2026. Written informed consent was obtained from the patients or, if a patient was unconscious, from their representatives. The study protocol was registered before the study commenced with the Chinese Clinical Trial Registry (ChiCTR; registration number: ChiCTR2300077594). All study procedures adhered to the principles of the Declaration of Helsinki for medical research involving human participants.

#### 2.1.1 Baseline Digital Twin Workflow

To establish a benchmark, we review the digital twin workflow proposed in [1]. Its forward prediction model uses a PatchTST architecture and a sliding-window strategy, taking the most recent 30 minutes of a patient’s glucose data as input to forecast glucose levels over a future horizon [2]. Performance is monitored in real time using the Ensemble Relative Error (ERE) [1]. A model update through full fine-tuning is triggered only when the ERE exceeds a predefined threshold (e.g., *ϵ* = 5%). If the error exceeds a critical upper bound (*δ* = 10%), the system suspends forecasting for safety [1]. Here, we investigate residual reinforcement learning as a strategy for correcting digital twin predictions at each time step.

#### 2.1.2 Digital Twin Workflow via Residual Reinforcement Learning

To improve dynamic updating, we propose a digital twin workflow that integrates a pretrained PatchTST time-series forecasting model with a reinforcement learning (RL) agent. The workflow supports real-time adaptation in glucose forecasting through a patient-specific residual-correction mechanism.

The system consists of two interacting modules:

- **The Foundation Model (Baseline):** A pretrained PatchTST model serves as the predictive backbone. Rather than using its multi-step output directly, we apply the model recursively: a one-step-ahead forecast is fed back into the input window to generate subsequent predictions.
- **The Correction Agent (A2C):** An Actor–Critic (A2C) agent acts as a dynamic controller. It uses recent glucose trends and prediction errors to add a residual correction to the baseline forecast, adapting the prediction to patient-specific dynamics.

#### 2.1.3 Baseline Foundation Model

Let *D*_*t*_ = {*y*_1_, *y*_2_, …, *y*_*t*_} denote the glucose sequence observed up to time *t*. At each time step, we construct a look-back window of length *L* (e.g., *L* = 30) as

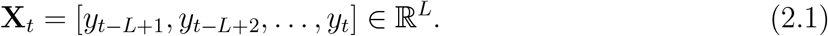

The frozen PatchTST baseline then produces a one-step-ahead forecast:

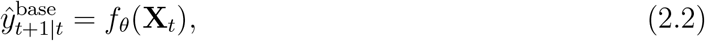

where *f*_*θ*_ denotes the pretrained PatchTST model with fixed parameters *θ*. While the pretrained model outputs a multi-step horizon (e.g., 15 minutes), we use only the first-step prediction as the baseline. Once the ground-truth observation *y*_*t*+1_ becomes available, we define the one-step residual as

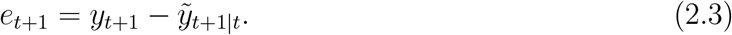

A history of recent residuals is maintained as **e**_*t*_ = [*e*_*t−L*+1_, …, *e*_*t*_] to inform the agent of short-term error trends.

#### 2.1.4. RL-Based Correction (A2C)

##### State Space

The RL state integrates the recent glucose trajectory and error history. Specifically, we define

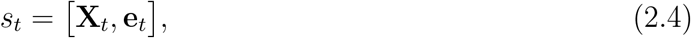

where **X**_*t*_ summarizes the current glucose dynamics and **e**_*t*_ captures recent patterns in the residuals.

##### Action Space and Policy

The RL agent employs a stochastic continuous policy during training, parameterized by an actor network with weights *ϕ*. Given the state *s*_*t*_, the actor outputs the mean *µ*_*ϕ*_(*s*_*t*_) and standard deviation *σ*_*ϕ*_(*s*_*t*_) of a Gaussian distribution, from which the raw action *a*_*t*_ is sampled:

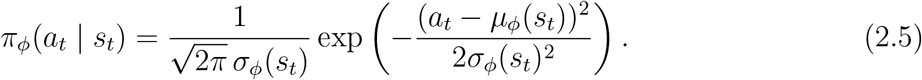

To ensure physiological safety and prevent excessively large corrections, the raw action is bounded before being applied to the glucose trajectory. Specifically, the final correction term is obtained by clipping *a*_*t*_ to the interval [*−c*_max_, *c*_max_]:

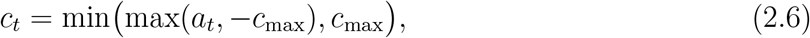

where *c*_max_ is a predefined hyperparameter representing the maximum allowable correction magnitude. In our experiments, we set *c*_max_ = 0.05.

During inference and deployment, we use the deterministic mean output of the actor, *a*_*t*_ = *µ*_*ϕ*_(*s*_*t*_). The bounded residual correction is then added to the baseline one-step-ahead forecast, yielding

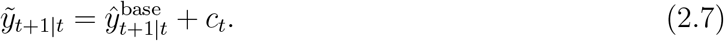

##### Reward Function

The reward *r*_*t*_ is designed to encourage accurate predictions while penalizing excessively large corrections. It is computed once the ground-truth observation *y*_*t*+1_ becomes available:

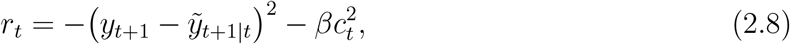

where the first term minimizes the squared error of the corrected prediction, and the second term regularizes the applied correction. Here, *β* is a regularization coefficient. In our experiments, we set *β* = 0.005.

##### Optimization Objective

The A2C agent consists of an actor *π*_*ϕ*_ and a critic *V*_*ω*_(*s*_*t*_), where *ω* denotes the parameters of the critic network. The overall objective is to maximize the expected cumulative return.

The critic is trained to estimate the state value function by minimizing the mean squared temporal-difference (TD) error:

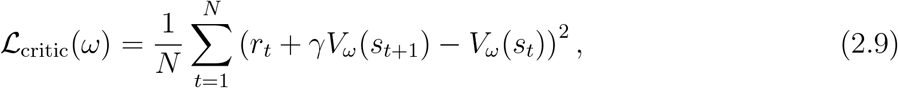

where *γ ∈* [0, 1) is the discount factor and *N* denotes the batch size.

The actor is updated via the policy gradient, where *A*_*t*_ = *r*_*t*_ + *γV*_*ω*_(*s*_*t*+1_) *− V*_*ω*_(*s*_*t*_) is used as an estimate of the advantage function. Accordingly, the actor loss is defined as

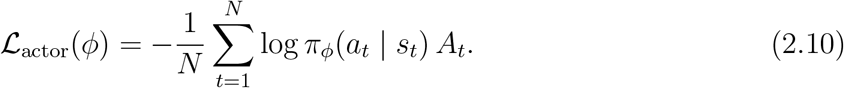

##### Multi-Step Forecasting

To generate an *H*-step forecast horizon 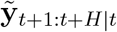 (with *H* = 15 in our experiments), we adopt a sliding-window strategy. The RL-based residual correction is applied only to the first prediction step in order to adjust the starting point of the forecast trajectory, while all subsequent steps are propagated using the frozen baseline dynamics.

Let 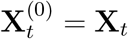. The first-step prediction is corrected by the RL agent as

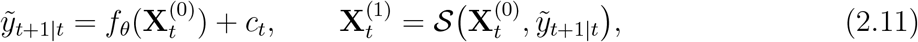

where *S*(*·, ·*) denotes the shift-and-append operator. Specifically, this operator removes the oldest entry in the current input window and appends the newly predicted value to the end, thereby preserving the fixed input length *L* required by the foundation model.

For *k* = 1, …, *H −* 1, the remaining forecast steps are generated recursively without further correction by the RL agent:

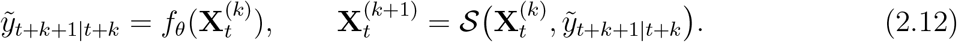

The resulting forecast sequence is given by

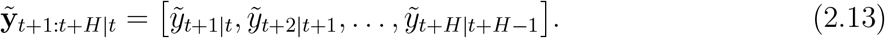

##### Algorithmic Workflow

###### Algorithm 1

Digital Twin RL-Correction: Training Loop

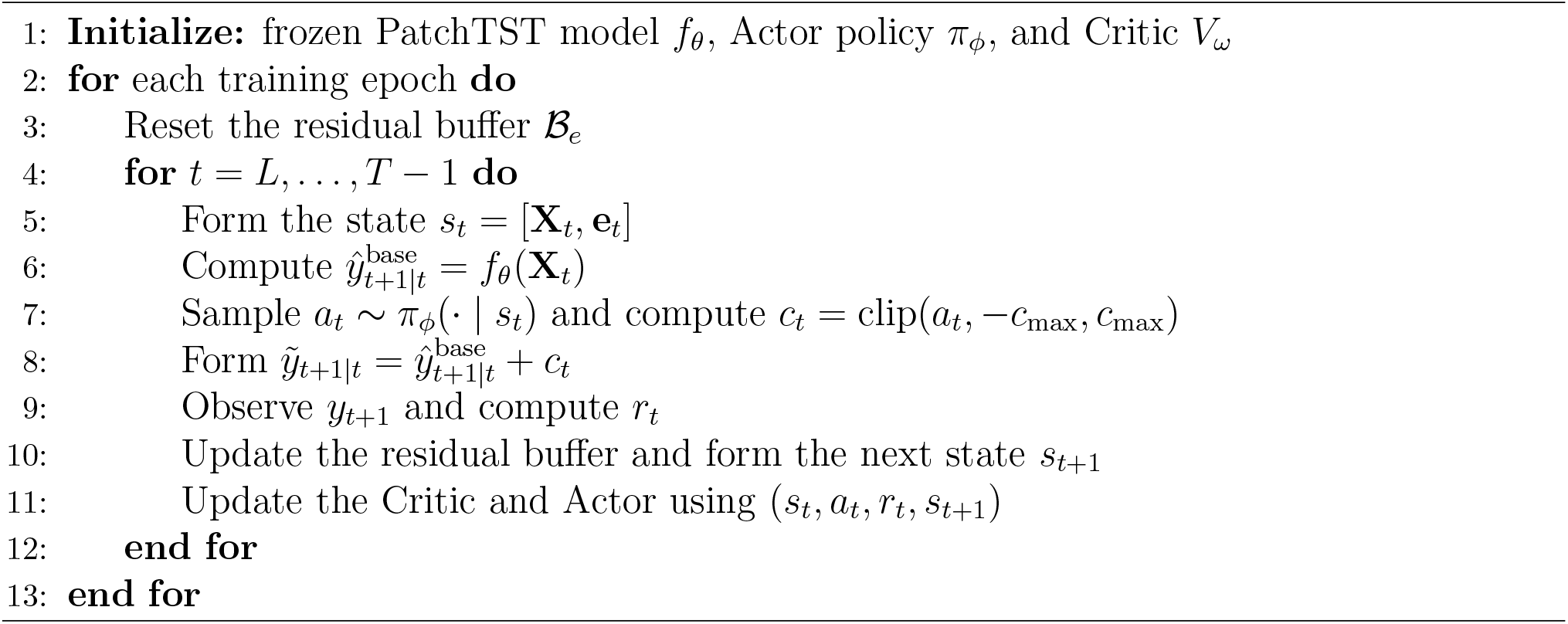

###### Algorithm 2

Digital Twin RL-Correction: Inference Loop

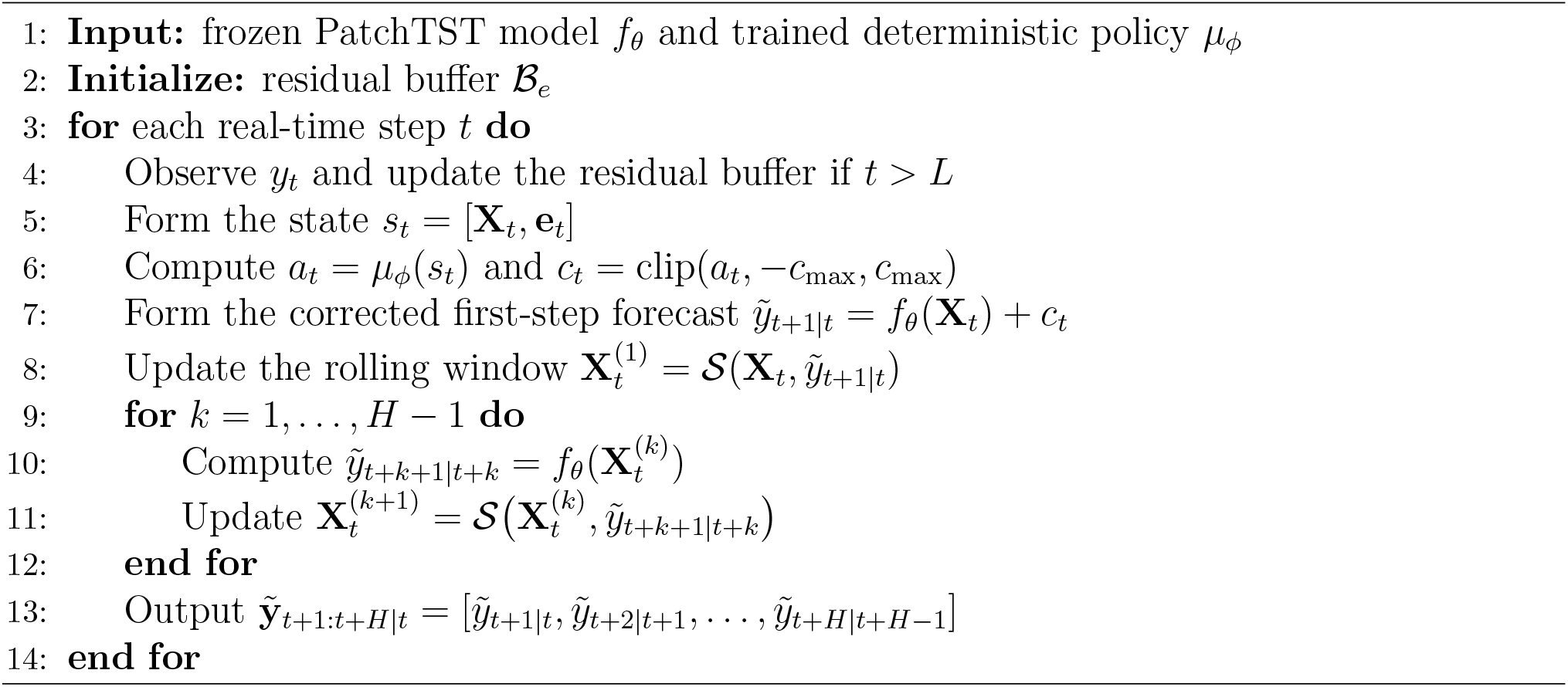

### 2.2 Hybrid Method

We combine the RL-based residual-correction framework with the previously developed transfer-learning workflow to construct a hybrid digital twin for real-time glucose monitoring and forecasting. The hybrid framework integrates continuous stepwise correction with threshold-triggered model adaptation. When a new patient is admitted, the pretrained foundation model is deployed immediately, and a pretrained RL agent applies a residual correction at each time step. The system also records the multi-step forecasts generated from each corrected first-step prediction. As in our previous workflow, we use a prediction horizon of *H* = 15 and the Ensemble Relative Error (ERE) to trigger model updates. The RL agent remains frozen throughout deployment, providing continuous online correction, while the base model is retrained as needed.

Figure 2.1 summarizes the proposed hybrid digital twin workflow, which consists of two data-assimilation loops. The inner loop performs local assimilation by correcting the one-step forecast at every prediction cycle. Its role is analogous to that of a Kalman filter: it adjusts the current forecast without directly modifying model parameters. The outer loop operates on a longer timescale and monitors forecasting error through the ensemble metric. When the base model diverges sufficiently from the evolving patient-specific dynamics that the RL controller can no longer compensate for the mismatch, the outer loop triggers retraining to reduce systematic bias and realign the digital twin with the patient’s trajectory. Thus, the hybrid method addresses both short-term prediction error and longer-term model bias. To formulate the hybrid framework, we first examine the observability of the forward prediction model.

**Figure 2.1:**
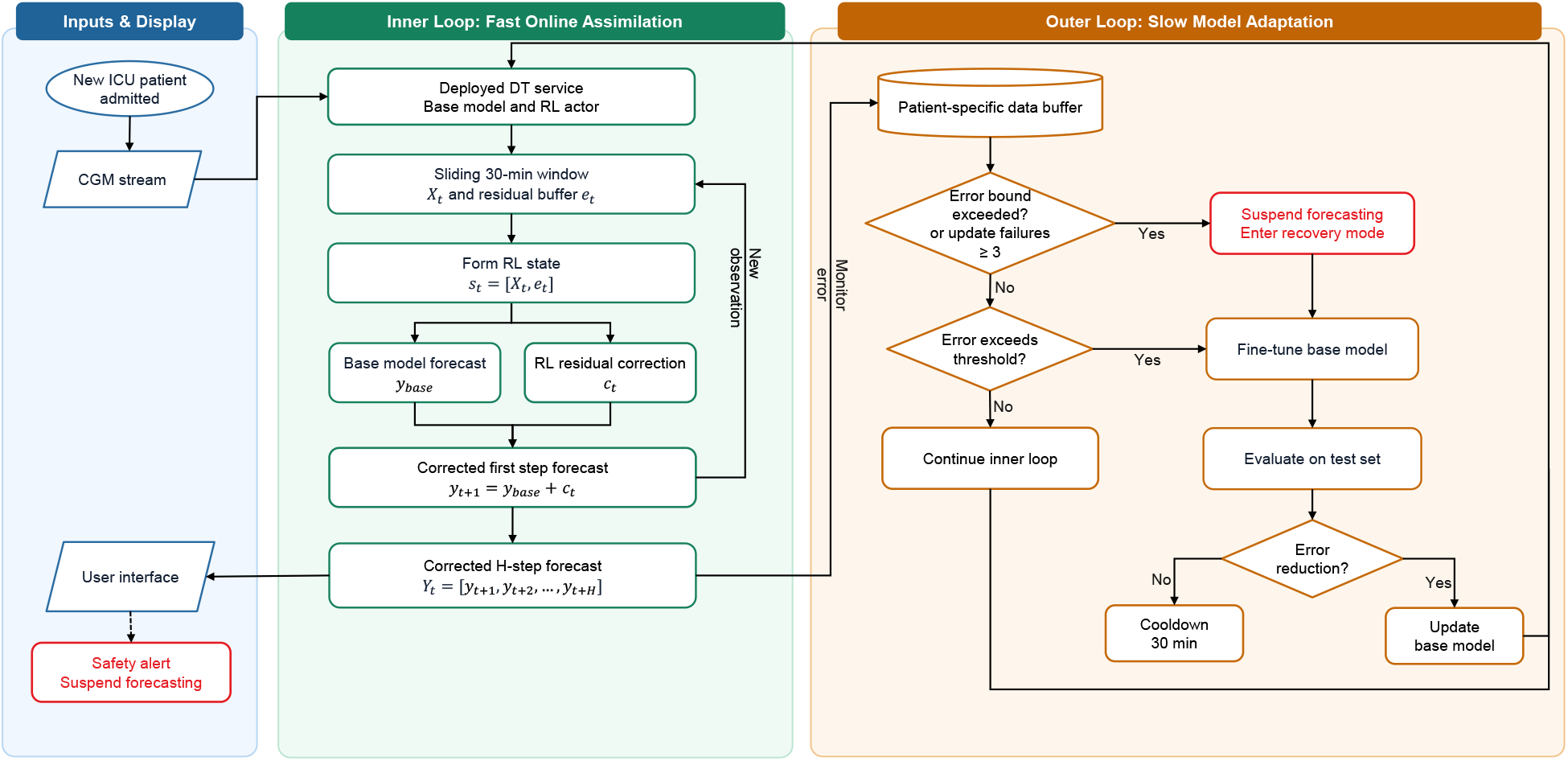
Schematic overview of the proposed hybrid digital twin workflow.

#### 2.2.1 Observability

Observability is a fundamental concept in Kalman filtering. Consider the linear discrete-time state-space model

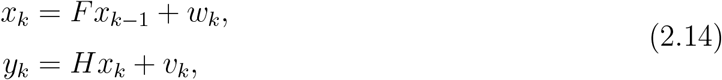

where *x*_*k*_ *∈* ℝ^*n*^ denotes the hidden state vector and *y*_*k*_ *∈* ℝ^*m*^ denotes the observation vector. To analyze observability, we focus on the deterministic part of the system, since observability is a structural property of the pair (*H, F*).

In the absence of noise, the state trajectory is completely determined by the initial state *x*_0_. Therefore, if *x*_0_ can be uniquely identified from the observations, then the entire state sequence can also be uniquely reconstructed. By stacking the observations over *n* consecutive time steps, we obtain

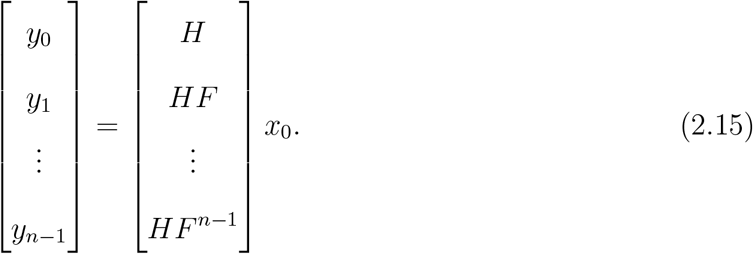

The matrix

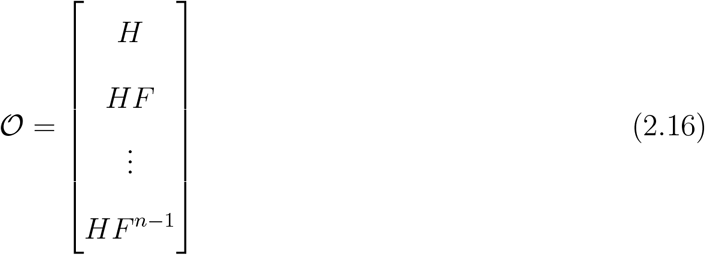

is called the observability matrix. The initial state *x*_0_ can be uniquely determined from the observation sequence if and only if *O* has full column rank, i.e.,

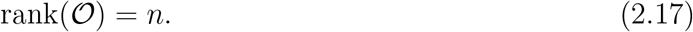

Under this condition, the system is said to be observable, and the full state trajectory can be uniquely recovered from the measurements.

#### 2.2.2 Nonlinear State-Space Formulation

Let

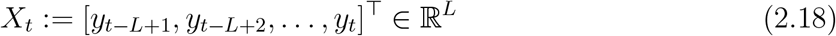

denote the observation window of length *L*. In our framework, the next observation is modeled as

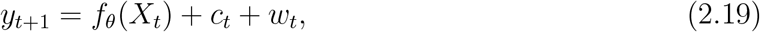

where *f*_*θ*_ is the frozen PatchTST predictor, *c*_*t*_ is the control input generated by the RL agent, and *w*_*t*_ denotes white noise.

Substituting the above expression into the shift-and-append operator yields

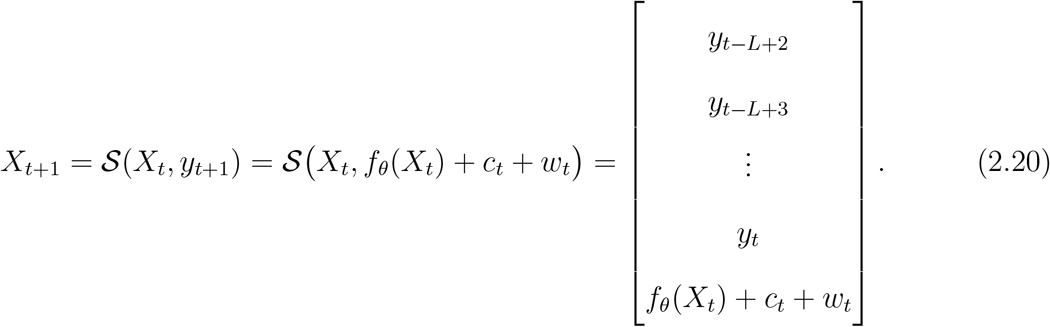

Thus, the state update removes the oldest entry and appends the newly generated value, while preserving the fixed window length *L*.

Equivalently, let *A*_*s*_ *∈* ℝ^*L×L*^ denote the shift matrix

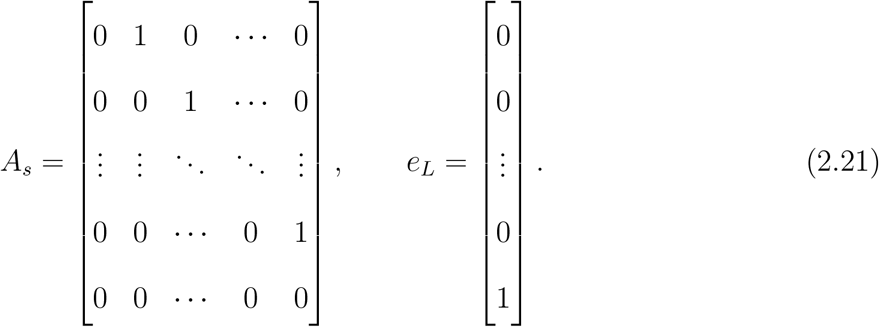

Then the state dynamics can be written compactly as

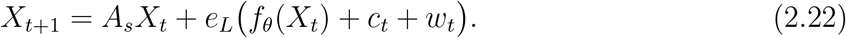

In this representation, the realized observation is simply the last coordinate of the updated state, namely,

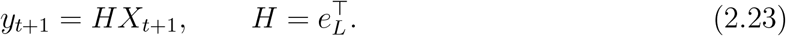

##### Local observability

To study observability, we consider the noise-free dynamics

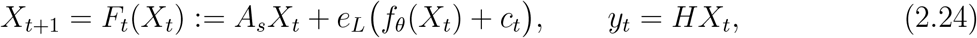

Since *f*_*θ*_ is nonlinear, we examine observability locally along a nominal trajectory 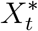.

Linearizing the dynamics around 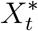 yields

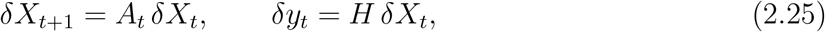

with Jacobian

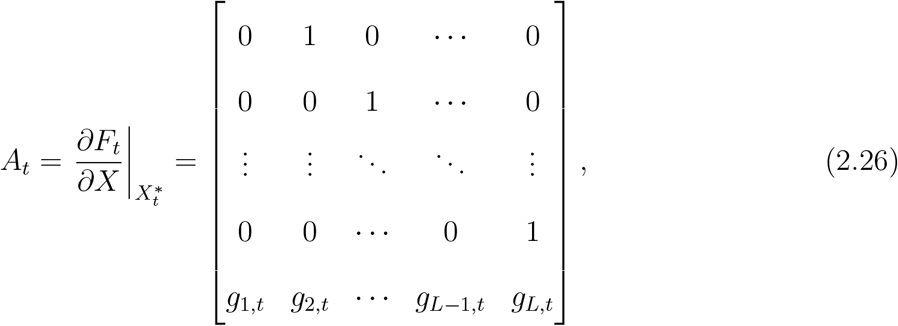

where

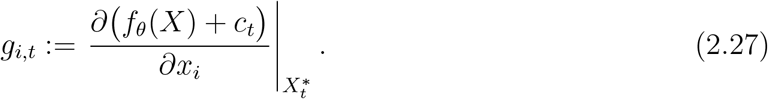

The corresponding *L*-step local observability matrix is given by

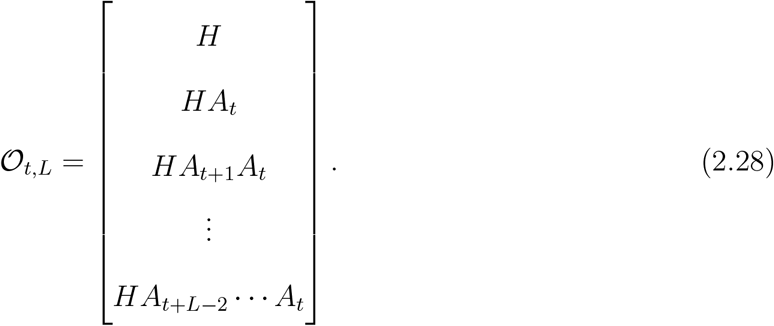

For the shift-and-append structure above, one obtains

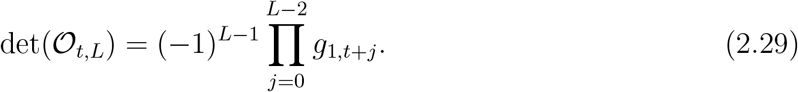

Therefore, the system is locally observable if and only if

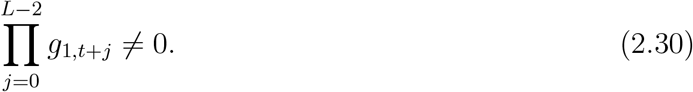

Equivalently, local observability is guaranteed provided that

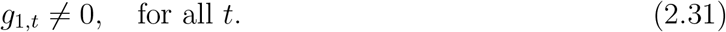

This condition indicates that the oldest component in the observation window must influence the newly generated observation at each step. Otherwise, its information is lost under the shift operation and cannot be recovered from future outputs.

#### 2.2.3 Inner Loop: RL-Based Correction

Let

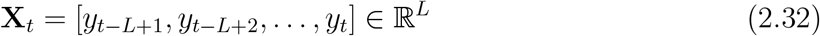

denote the current look-back window, where *L* = 30. At time *t*, the current base model 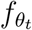 produces a one-step baseline forecast

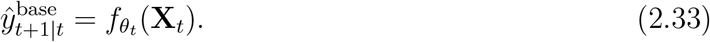

To perform real-time correction, we define the RL state as

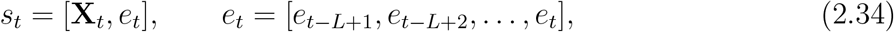

where *e*_*t*_ denotes the recent residual history. During deployment, the pretrained actor network with parameters *ϕ* outputs the deterministic mean action *µ*_*ϕ*_(*s*_*t*_). The bounded correction is then given by

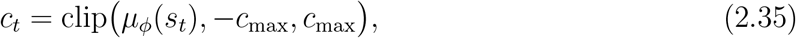

where *c*_max_ *>* 0 is the maximum allowable correction magnitude. The corrected first-step forecast is

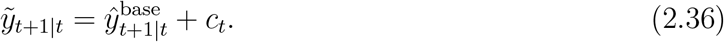

Once the observation *y*_*t*+1_ becomes available, the corrected one-step residual is computed as

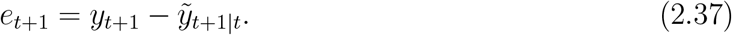

##### Multi-Step Forecasting

To generate a forecast horizon of length *H* = 15, we use the corrected first-step forecast as the starting point of a recursive rollout. Let

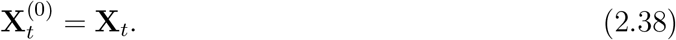

The first step is corrected by the RL controller:

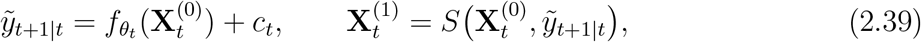

where *S*(*·, ·*) denotes the shift-and-append operator.

For *k* = 1, …, *H −* 1, the remaining forecast steps are generated recursively by the base model without further RL correction:

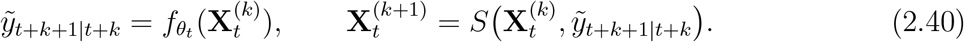

Thus, the complete forecast sequence is

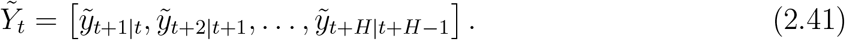

#### 2.2.4 Outer Loop: Base-Model Updating

To monitor performance in real time, we retain the ensemble mechanism from the previous digital twin workflow. At each prediction time *i*, the hybrid twin produces a corrected *H*-step forecast

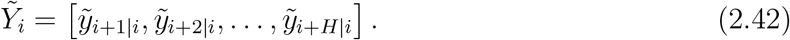

Because consecutive rolling windows overlap, multiple forecasts generated at earlier times predict the same target time *t*. We therefore define the set of overlapping corrected predictions as

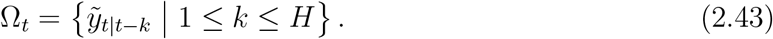

Following the previous workflow, these overlapping forecasts are combined using exponentially decaying weights

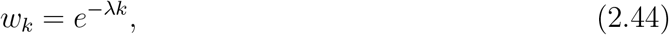

where *λ ≥* 0 is a decay-rate hyperparameter. The ensemble prediction at target time *t* is then given by

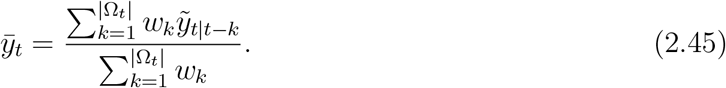

The corresponding Ensemble Relative Error (ERE) is defined by ERE_*t*_

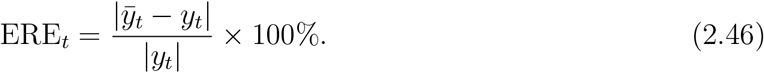

In the hybrid workflow, ERE_*t*_ serves as the primary monitoring signal for potential model updating. Let *ϵ* denote the update threshold and *δ* denote the safety stop threshold:

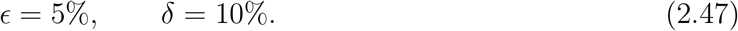

If ERE_*t*_ *≥ ϵ*, the system triggers a base-model updating procedure using the accumulated patient-specific data. If ERE_*t*_ *≥ δ*, forecast output and digital twin operation are suspended for safety.

When an update is triggered, the accumulated patient-specific samples are partitioned into temporary training, validation, and test subsets. A candidate updated base model is then obtained by fine-tuning the current base model on the new patient data. Before redeployment, the candidate model is compared with the current base model on the temporary test set, and the candidate is accepted only if it achieves better predictive performance. Otherwise, the current model is retained.

If an update attempt fails, the system enters a cooldown period to accumulate additional patient data before another update is attempted. After repeated failures, forecasting remains suspended, and manual review is required.

Once a pause is triggered at time *τ*_*p*_, forecasting output is suspended immediately, while raw CGM acquisition continues. The currently deployed base model 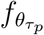 and all patient-specific data accumulated up to *τ*_*p*_ are retained for recovery. Automatic restart is not attempted immediately after a pause. Instead, the system enters a recovery mode and accumulates a recovery window of length *L*. These new glucose observations are used to reconstruct the glucose window *X*_*t*_, and the residual buffer *e*_*t*_ is reinitialized to zero, restoring the hybrid state for restart evaluation.

During recovery, we consider a candidate set of restart models based on the fine-tuning strategies developed in [1], including (i) linear probing, (ii) full fine-tuning, and (iii) staged fine-tuning. Each candidate is trained using the same training and validation sets and evaluated on the same test set. Consistent with the update rule, a candidate is accepted only if it achieves better performance than 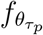 on the test set. Otherwise, 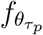 is retained as the restart model. Among all eligible candidates, the one with the best performance is selected for restart.

After a restart model has been selected, the system does not immediately return to full online deployment. Instead, the selected model is first run in recovery mode over a prescribed observation interval. If the ERE remains below the update threshold *ϵ* = 5% throughout this interval, the model is redeployed online. Otherwise, the system remains in recovery mode, accumulates additional patient-specific data, and continues fine-tuning candidate restart models.

## 3 Results

### 3.1 Model with RL-Based Correction

#### 3.1.1 Pretraining of the RL Agent

We first pretrain the RL agent on the source training dataset. Since no historical prediction errors are available at the beginning of the sequence, all residuals are initialized to zero. When the first ground-truth observation becomes available at time step *t* = 31, the first observed residual is calculated, replacing the most recent zero-initialized entry in the residual state. The agent is trained for 20 epochs on the training set, and the final model is selected based on the highest cumulative reward achieved on the validation set.

The cumulative reward trajectories over training epochs on the training and validation sets are shown in Fig. 3.1. The learning curves exhibit a steady upward trend during training, with the best validation performance achieved at epoch 8. The model from this epoch was then evaluated on the test set. In this experiment, a 30-step look-back window was used to predict the next glucose value. Specifically, given the input window *X*_*t*_ = [*y*_*t−*29_, …, *y*_*t*_], the model generated a one-step-ahead prediction *ŷ*_*t*+1|*t*_. The one-step absolute percentage error (APE) was defined as

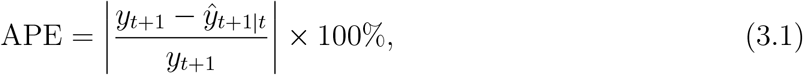

where *y*_*t*+1_ is the observed glucose value at the next time point and *ŷ*_*t*+1|*t*_ is the corresponding one-step prediction made at time *t*. The reported MAPE was then computed by averaging APE over the entire test trajectory. After applying the RL-based correction, the MAPE decreased from 0.49% to 0.34%, indicating a substantial improvement in one-step forecasting accuracy.

**Figure 3.1:**
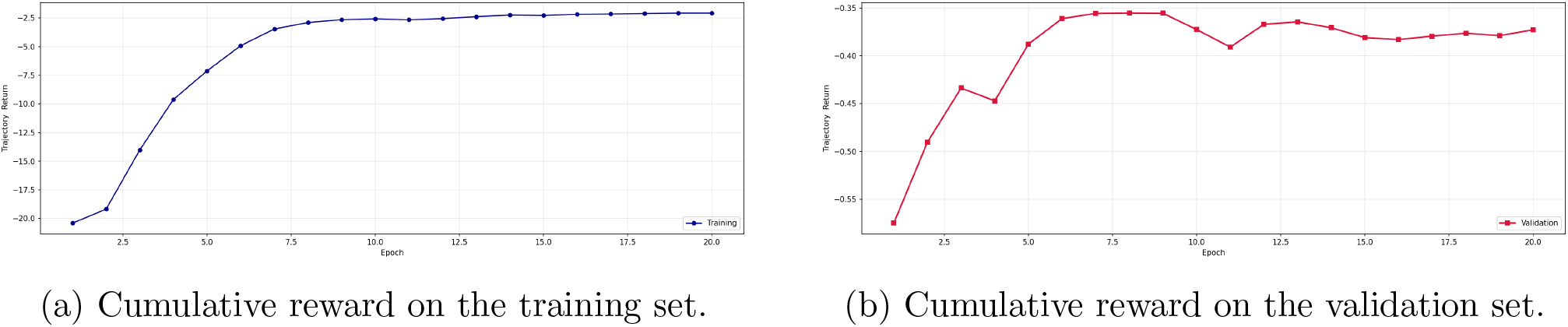
Cumulative reward trajectories of the RL agent during pretraining on the training and validation sets across 20 epochs.

#### 3.1.2 Evaluation on New Patients

We next evaluate the pretrained RL agent on new patients without any further adaptation. As shown in Table 3.1, the RL-based correction consistently improves forecasting performance across all 10 patients, leading to lower MAPE values than the baseline model in every case. This indicates that the pretrained correction policy generalizes well to unseen patients and can effectively reduce predictive error without patient-specific retraining.

Figure 3.2 further illustrates the performance improvement: the gray curve shows the baseline prediction error, and the blue curve shows the error after RL-based correction. The blue curve generally remains below the gray curve, indicating lower errors for most of the observation period. Figure 3.3 presents the corrections generated by the RL agent during deployment. All corrections remain within the predefined range, confirming that the policy respects the imposed bound while adjusting the baseline forecasts.

**Figure 3.2:**
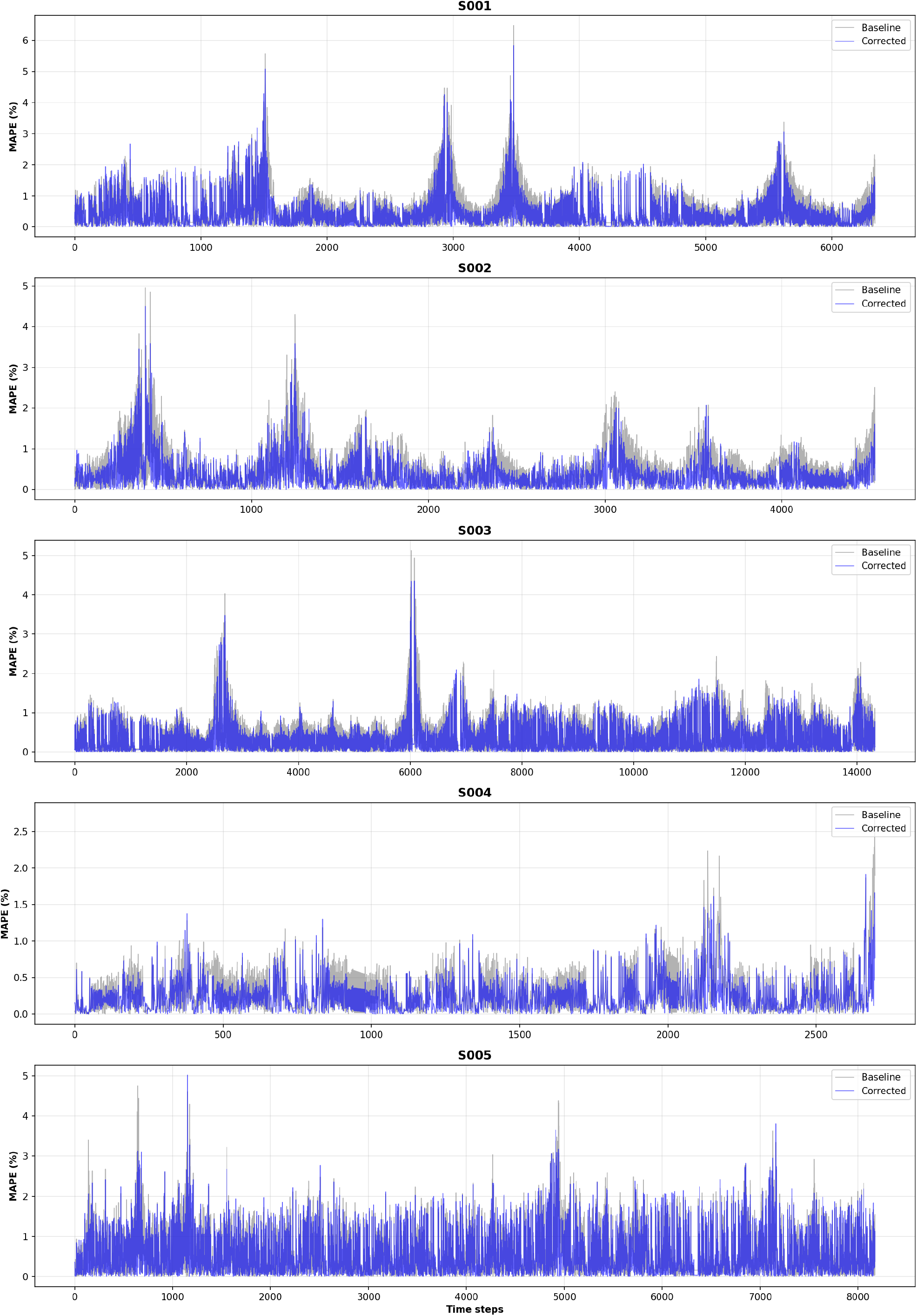
Comparison of prediction errors for patients S001–S005. The gray curves show errors from the baseline forecasts, and the blue curves show errors after RL-based correction. Corrected errors remain below 5% except for two slight exceedances in S001 and one in S005. The corrected errors remain lower than the uncorrected errors throughout the observation 16 periods, illustrating the improvement provided by RL correction.

**Figure 3.3:**
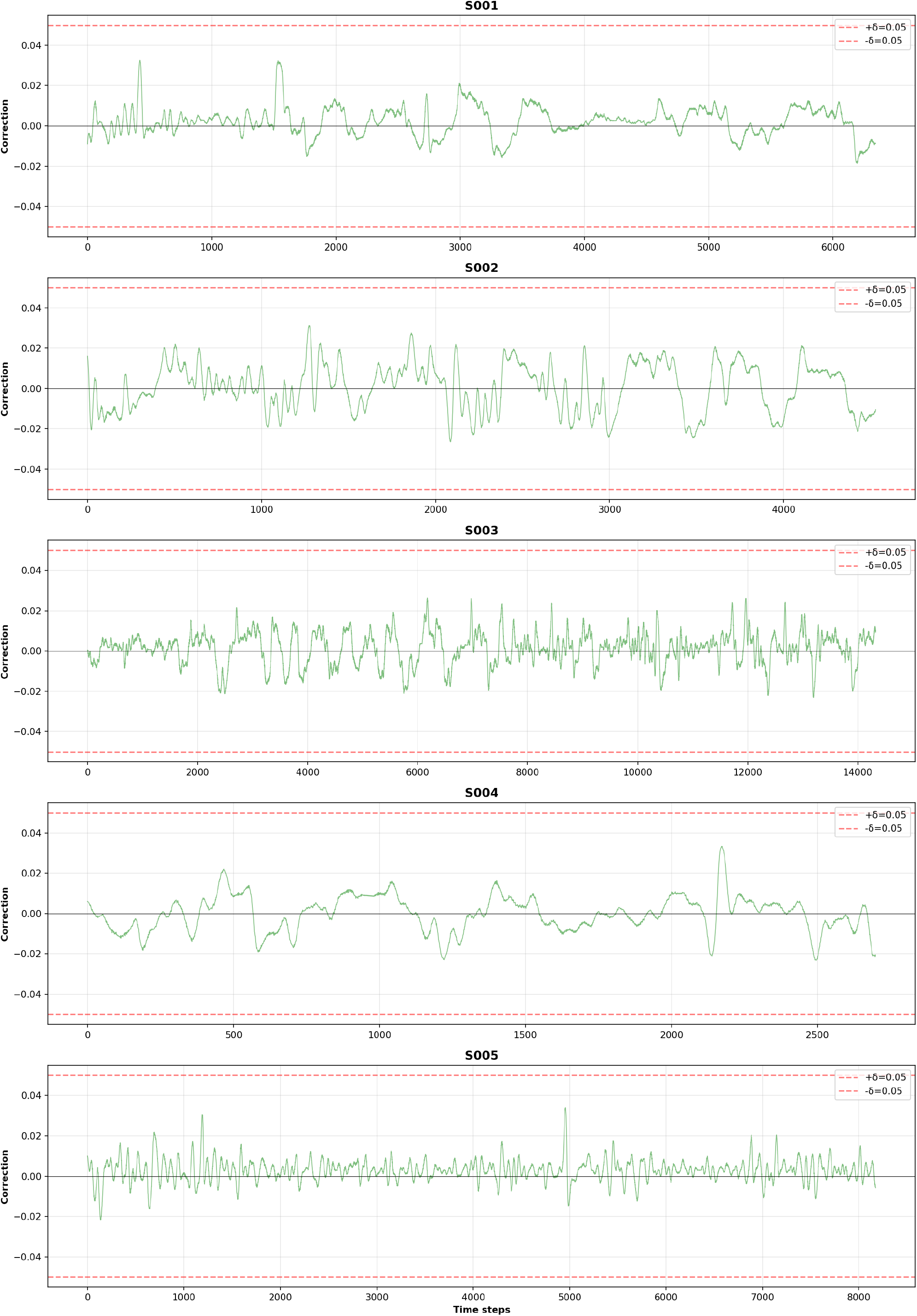
Bounded corrections generated by the RL agent during deployment for patients S001–S005 over the observation intervals. Corrections remain within the prescribed range of (*−*0.05, 0.05).

These results show that RL-based correction improves the baseline model’s ability to forecast individual glucose trajectories, with relative errors generally *<* 5%. Exceedances of the predefined 5% error tolerance are rare in the evaluated trajectories, suggesting that this approach may support patient-specific forecasting without dynamic model updates. Because the previous digital twin workflow is reactive whereas RL-based correction provides proactive forecast adjustment, we next combine the two approaches in a hybrid workflow that forecasts future trajectories and updates the model as needed.

### 3.2 hybrid DT Model

The corresponding results of the hybrid DT model are summarized in Table 3.2, which reports the trajectory-averaged ERE and the maximum ERE for both the original DT and the proposed hybrid DT. As shown in the table, the hybrid DT achieves a lower mean ERE in nine of the ten patients, with relative improvements ranging from 1.27% to 9.76%. Although the mean ERE for patient S005 increases slightly, the hybrid DT reduces the maximum ERE for all ten patients. This indicates that, in addition to improving average performance in most cases, the hybrid method is particularly effective in reducing large predictive errors.

**Table 3.2:** Comparison of trajectory-averaged and maximum ERE between the original DT and the proposed hybrid DT for patients S001–S010.

| Patient | DT (%) | Hybrid DT (%) | Improvement (%) | DT Max (%) | Hybrid DT Max (%) |
| --- | --- | --- | --- | --- | --- |
| S001 | 0.520 | 0.496 | 4.56 | 7.34 | 6.72 |
| S002 | 0.470 | 0.424 | 9.76 | 5.00 | 4.68 |
| S003 | 0.339 | 0.317 | 6.39 | 4.98 | 4.52 |
| S004 | 0.283 | 0.267 | 5.72 | 2.08 | 2.00 |
| S005 | 0.584 | 0.607 | -3.84 | 6.20 | 5.74 |
| S006 | 0.322 | 0.306 | 4.77 | 2.64 | 2.42 |
| S007 | 0.259 | 0.257 | 0.87 | 1.73 | 1.69 |
| S008 | 0.324 | 0.295 | 8.77 | 1.74 | 1.63 |
| S009 | 0.306 | 0.293 | 4.24 | 1.54 | 1.41 |
| S010 | 0.245 | 0.235 | 3.77 | 1.98 | 1.91 |

Figure 3.4 further illustrates this comparison. The gray curves show the ERE of the original DT, and the blue curves show the ERE of the hybrid DT. The blue curves generally remain below the gray curves, with particularly clear improvements near error peaks. These results suggest that the hybrid workflow reduces extreme forecasting errors while maintaining competitive average accuracy.

**Figure 3.4:**
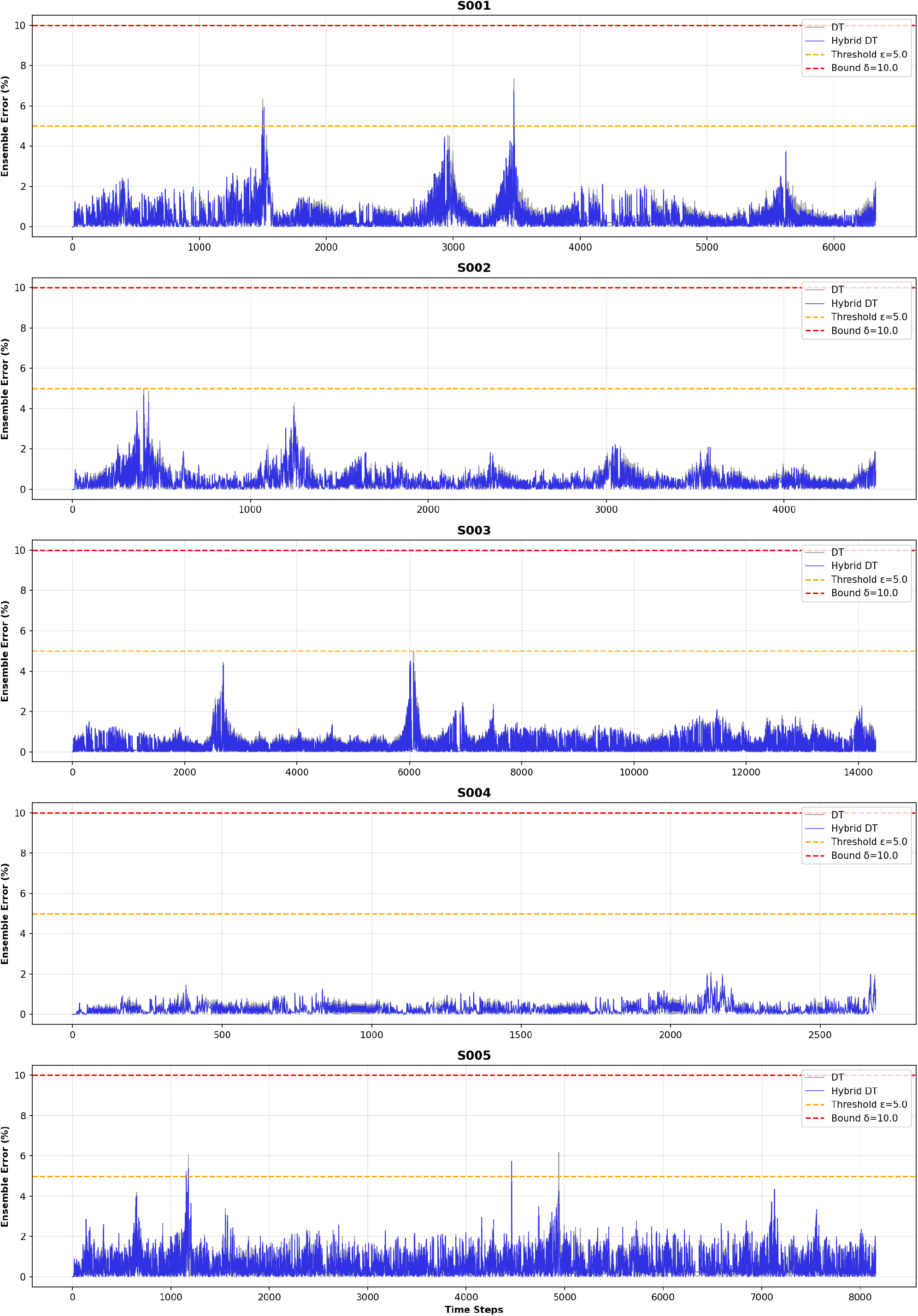
Comparison of the ERE for patients S001–S005. The gray curves denote results from the original DT, while the blue curves denote those from the proposed hybrid DT.

#### 3.2.1 Two Timescales

The hybrid digital twin can be viewed as a two-timescale adaptive system:

1. **Inner loop (short timescale)**. At each time step, the RL agent adjusts the one-step forecast. This loop acts locally and continuously to compensate for short-term forecasting errors, but does not update model parameters.
2. **Outer loop (long timescale)**. The ensemble error is monitored across overlapping multi-step forecasts. When persistent mismatch indicates structural drift that cannot be compensated by the RL correction, the base model is retrained and redeployed.

This design ensures that the inner-loop controller and the outer-loop updating mechanism play distinct and complementary roles, yielding a hybrid digital twin that is both continuously adaptive and operationally stable.

Mathematically, the method may also be interpreted as a two-timescale optimization scheme. The state evolves on a short timescale and is updated at every time step, whereas the model parameters evolve on a longer timescale and are updated only when the ensemble error becomes sufficiently large. This interpretation clarifies the roles of the two loops: the inner loop addresses local forecast error, while the outer loop adapts the forecasting model to longer-term changes.

#### 3.2.2 Alternative Trigger Metric

We also consider an alternative trigger metric for model updating. Unlike the previous trigger, which is based on the ensemble-averaged error, this metric is based on the number of overlapping predictions whose individual relative errors exceed predefined thresholds.

Recall that, for a target time point *t*, the set of overlapping corrected predictions is

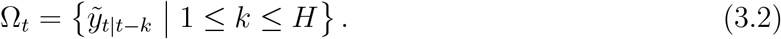

For each 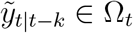, we define its individual relative error by

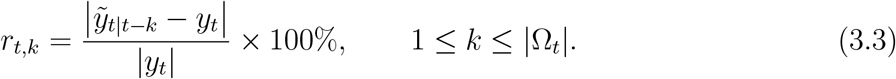

We then introduce the count-based update statistic

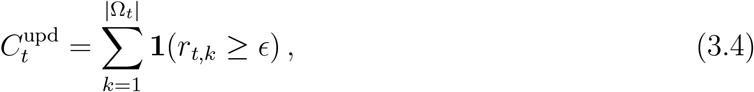

where **1**(*·*) denotes the indicator function and *ϵ* = 5% is the update threshold. In this study, the fine-tuning process is triggered whenever

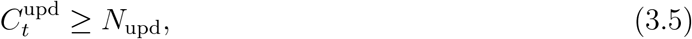

where *N*_upd_ = 12.

Similarly, we define the count-based safety statistic

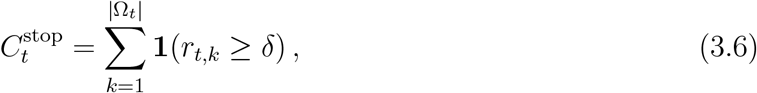

where *δ* = 10% is the safety threshold. The forecasting workflow is suspended whenever

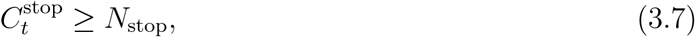

where *N*_stop_ = 12 in the present study. All other safety and updating procedures remain the same as those described previously.

The corresponding results are shown in Fig. 3.5. Compared with the previous workflow, the count-based trigger leads to more frequent model updates. In the previous workflow, the trigger metric was based on the ensemble-averaged error with exponentially decaying weights, so that the shortest-horizon prediction, in particular the first-step prediction, received the largest weight. In contrast, the present count-based criterion reflects how many predictions exceed the predefined error thresholds, and is therefore more sensitive to error accumulation across the recursive rollout. As a result, the fine-tuning procedure is triggered more frequently than in the previous workflow.

**Figure 3.5:**
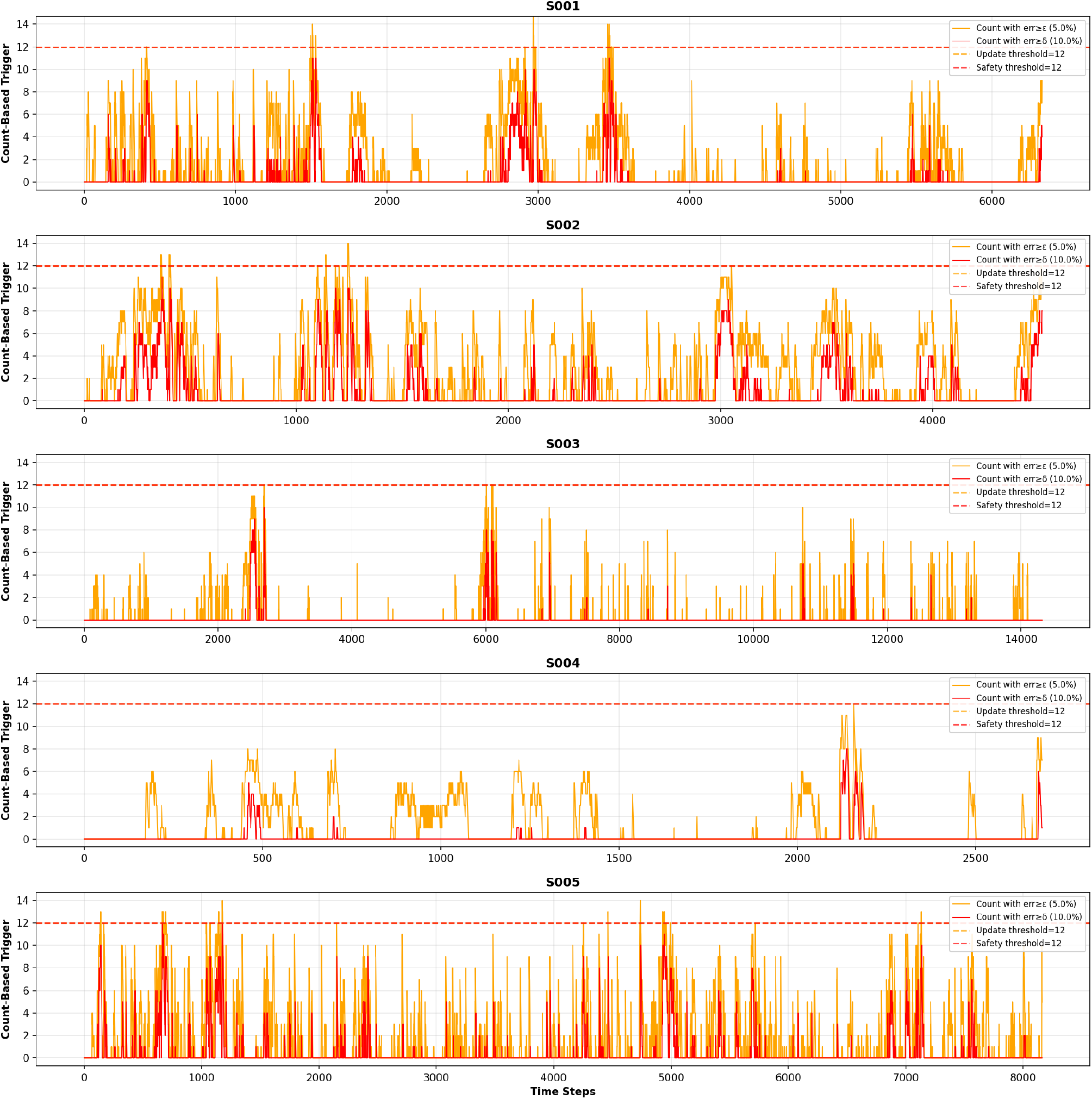
Count-based trigger statistics for patients S001–S005. The orange curves denote the number of predictions with relative errors exceeding 5%, while the red curves denote the number exceeding 10%. The dashed lines indicate the corresponding update and safety count thresholds.

### 3.3 Comparison with the RL-only DT

To further evaluate the contribution of the RL-based residual correction module, we constructed an RL-only model. In this setting, the DT backbone was kept fixed during deployment, and only the RL agent was used to perform stepwise residual correction. Therefore, unlike the proposed hybrid model, the RL-only model did not include threshold-triggered fine-tuning or outer-loop backbone updating. This comparison allows us to examine whether the RL correction module alone can improve DT predictions without dynamically updating the backbone model. The trajectory-averaged ERE and maximum ERE of the original DT and the RL-only model are reported in Table 3.3.

**Table 3.3:** Comparison of trajectory-averaged ERE and maximum ERE between the original DT and the RL-only model for patients S001–S010.

| Patient | DT (%) | RL-only (%) | Improvement (%) | DT Max (%) | RL-only Max (%) |
| --- | --- | --- | --- | --- | --- |
| S001 | 0.520 | 0.468 | 10.02 | 7.34 | 6.79 |
| S002 | 0.470 | 0.424 | 9.76 | 5.00 | 4.68 |
| S003 | 0.339 | 0.317 | 6.39 | 4.98 | 4.52 |
| S004 | 0.283 | 0.267 | 5.72 | 2.08 | 2.00 |
| S005 | 0.584 | 0.563 | 3.56 | 6.20 | 5.05 |
| S006 | 0.322 | 0.306 | 4.77 | 2.64 | 2.42 |
| S007 | 0.259 | 0.257 | 0.87 | 1.73 | 1.69 |
| S008 | 0.324 | 0.295 | 8.77 | 1.74 | 1.63 |
| S009 | 0.306 | 0.293 | 4.24 | 1.54 | 1.41 |
| S010 | 0.245 | 0.235 | 3.77 | 1.98 | 1.91 |

Compared with the original DT, the RL-only model reduced the trajectory-averaged ERE for all ten patients, with relative improvements ranging from 0.87% to 10.02%. The maximum ERE was also reduced in every patient. These results suggest that RL-based residual correction alone can improve prediction accuracy.

This comparison is further illustrated in Fig. 3.6, where the gray curves represent the ERE of the original DT and the blue curves represent the ERE of the RL-only DT. Across the representative patients, the RL-only model generally reduced local error excursions and lowered several high-error peaks, while keeping the ERE below the 10% safety bound. Consistent with the quantitative results in Table 3.3, the RL-only model achieved lower maximum ERE than the original DT.

**Figure 3.6:**
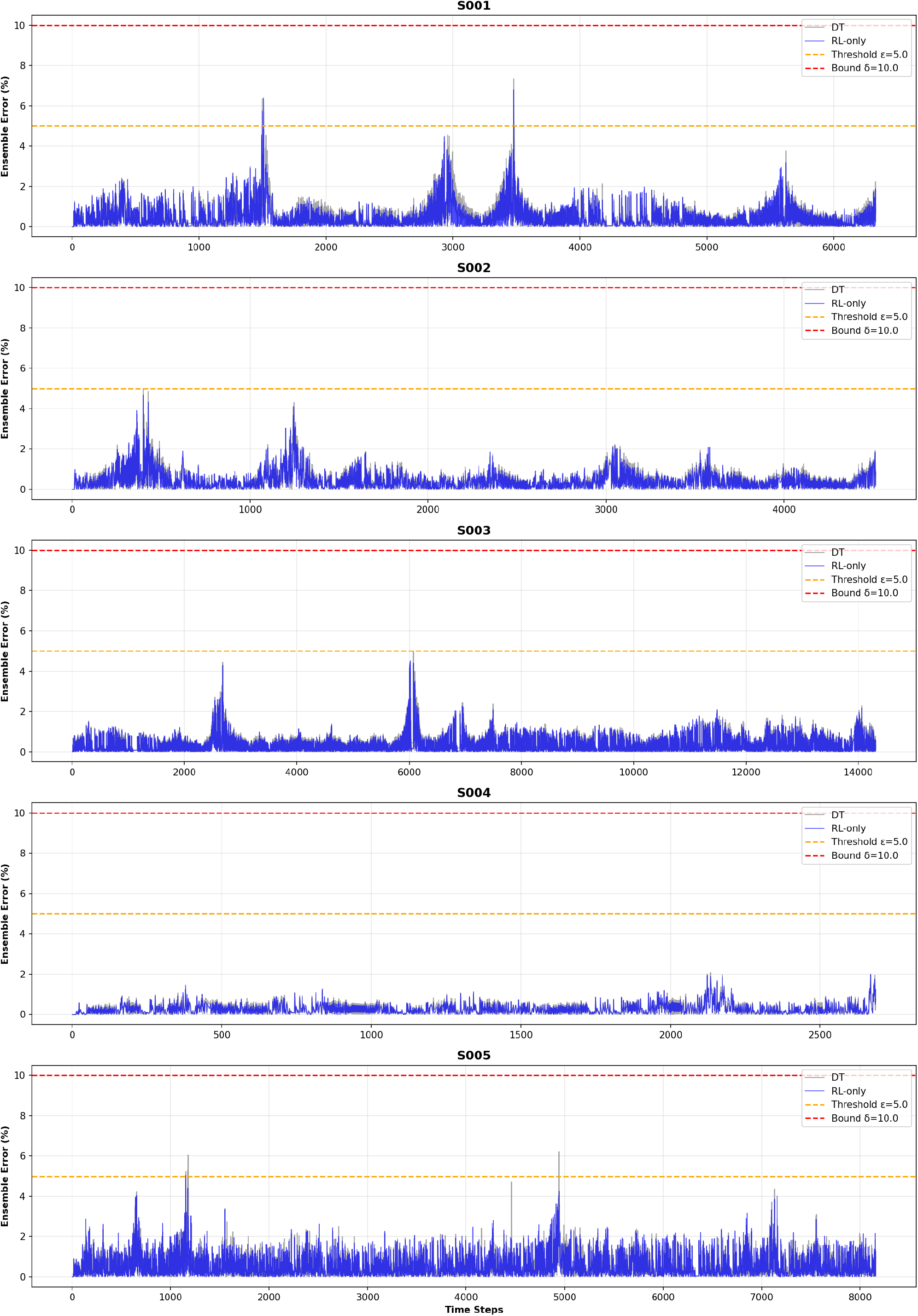
Comparison of the ERE for patients S001–S005. The gray curves denote results from the original DT, while the blue curves denote those from the RL-only DT.

Only patients S001 and S005 triggered the outer-loop fine-tuning procedure. For the remaining patients, the hybrid DT and the RL-only DT were therefore expected to produce the same results, because the backbone model was not updated. However, in patients S001 and S005, the fine-tuning step did not yield additional improvement beyond RL-only correction. In S005, it even led to worse performance. This observation suggests that most of the performance gain in the current hybrid workflow may be attributable to the RL residual correction module rather than to the outer-loop backbone updating mechanism. It also motivates reconsideration of the current fine-tuning strategy and trigger criterion.

One possible explanation is the interaction between backbone updating and the learned RL policy. The RL agent was trained to correct the residual pattern generated by the original frozen DT backbone. Once the backbone is fine-tuned, the residual distribution observed by the RL agent may shift. This policy–backbone mismatch may reduce the effectiveness of the learned correction policy and may even lead to slight degradation in some patients. Therefore, although the hybrid DT provides a conceptually useful two-timescale framework, its current outer-loop updating strategy may require additional safeguards.

These findings suggest that the RL-only DT can serve as a lightweight and effective residual-assimilation strategy. Compared with the hybrid DT, the RL-only model avoids repeated backbone fine-tuning, reduces computational complexity, and preserves a stable residual distribution for the RL agent. The hybrid DT may still be useful when persistent structural drift occurs, but future implementations should consider a more selective trigger metric and a refined fine-tuning strategy.

## 4 Discussion

In this study, we develop a proactive digital twin workflow for continuous glucose forecasting by coupling a pretrained PatchTST foundation model with an Actor–Critic RL agent that provides bounded residual corrections in real time. The motivation is to address a limitation of our previous transfer-learning workflow: although threshold-triggered fine-tuning can restore performance after substantial drift, it remains reactive and offers limited support for continuous data assimilation. The proposed residual RL framework instead adjusts the forecast at each prediction cycle, allowing the digital twin to respond to short-term discrepancies between baseline predictions and the observed glucose trajectory.

Several findings support the value of this formulation. First, pretraining on the source dataset produced a stable learning process, with cumulative reward increasing on both the training and validation sets and the best validation performance achieved by epoch 8. Second, on the held-out source test set, RL correction reduced the MAPE from 0.49% to 0.34%, showing that even when the foundation model is already accurate, a learned residual controller can further improve one-step forecasting performance. Third, the pretrained agent generalized to all 10 new patients without additional patient-specific retraining. As summarized in Table 3.1, every patient exhibited improvement after correction, with relative gains ranging from modest but consistent reductions (for example, S005 and S007) to substantial error decreases exceeding 25% for S002 and 30% for S008. This uniform direction of improvement suggests that the RL agent learned a transferable correction policy rather than merely overfitting to idiosyncrasies of the source patient.

**Table 3.1:** MAPE comparison between the baseline forecasts and the RL-corrected forecasts across all 10 patients.

| Patient | Baseline MAPE (%) | Corrected MAPE (%) | Relative Improvement (%) |
| --- | --- | --- | --- |
| S001 | 0.48 | 0.43 | 10.5 |
| S002 | 0.49 | 0.36 | 26.73 |
| S003 | 0.34 | 0.29 | 13.84 |
| S004 | 0.29 | 0.25 | 13.84 |
| S005 | 0.54 | 0.52 | 3.90 |
| S006 | 0.32 | 0.27 | 15.88 |
| S007 | 0.26 | 0.25 | 3.80 |
| S008 | 0.35 | 0.24 | 30.92 |
| S009 | 0.30 | 0.28 | 8.49 |
| S010 | 0.24 | 0.23 | 6.19 |

The proposed workflow is attractive because it separates prediction and adaptation into complementary roles. The pretrained PatchTST serves as a strong global prior that captures the dominant temporal structure of glucose dynamics, while the RL module acts locally to compensate for short-term mismatch. By keeping the backbone frozen, inference remains computationally light and avoids repeated fine-tuning, which can be costly and unstable in a real-time ICU setting. At the same time, the RL correction operates only on the first forecast step, thereby shifting the initial condition of the predictive trajectory without repeatedly perturbing all subsequent forecasted points. This design preserves the intrinsic temporal dynamics learned by the foundation model while still enabling immediate assimilation of recent patient-specific information.

The qualitative results are also informative. The prediction-error comparisons in Fig. 3.2 show that the corrected forecasts remain below the baseline error curve for most of the deployment period across patients S001–S005, indicating sustained benefit rather than isolated improvements at a few time points. In parallel, Fig. 3.3 shows that the learned corrections remain well within the prescribed bounds of *±δ* = 0.05. This behavior is important for two reasons. From a modeling perspective, it demonstrates that the policy does not rely on erratic or excessively large interventions to achieve lower error. From a safety perspective, bounded correction limits the magnitude of forecast shifts in response to transient discrepancies. The regularization term in the reward function and the clipping operation in the action mapping together therefore appear to provide an effective mechanism for balancing responsiveness and stability.

A further conceptual contribution of this work is the use of reinforcement learning for data assimilation rather than classical control alone. In this setting, the RL state combines the recent glucose trajectory and recent residual history, so the agent learns not only the current glucose level and trend but also the recent pattern of model misfit. This makes the correction policy adaptive to both physiological evolution and forecasting bias. Such a strategy is especially relevant in critically ill patients, where abrupt metabolic changes, evolving treatment responses, and inter-patient variability may cause the relationship between past glucose values and near-future behavior to shift over time.

At the same time, the present study has several limitations. The most important is the small cohort size. Although the pretrained RL agent improved performance for all 10 evaluation patients, the cohort remains too limited to establish robustness across the full spectrum of ICU populations, levels of illness severity, treatment protocols, and glucose variability patterns. Broader external validation will be needed to determine how well the learned correction strategy transfers across institutions, sensor platforms, and patient subgroups. In addition, the source foundation model and RL policy were trained using glucose data alone. This choice was motivated partly by prior evidence that insulin data had limited impact on forecasting performance in the setting considered here, but it also means that the current digital twin does not explicitly incorporate other clinically meaningful covariates such as nutrition delivery, vasopressor use, corticosteroids, inflammatory markers, or broader physiological monitoring signals.

Another limitation concerns the forecasting architecture itself. In the present design, the RL agent corrects only the first prediction step, and the remaining *H −* 1 steps are propagated recursively using the frozen baseline model. This was a deliberate choice to maintain stability and interpretability, but it may also limit the extent to which the workflow can address error accumulation deeper in the forecast horizon. Future work could examine multi-step residual control, horizon-aware reward design, or hybrid formulations in which the agent decides when and where along the forecast trajectory a correction is most beneficial. It will also be important to investigate whether alternative RL methods, such as deterministic policy gradient methods or uncertainty-aware policies, offer better sample efficiency or more robust adaptation than the A2C formulation used here.

The study also focuses on predictive accuracy, quantified primarily by MAPE and cumulative reward. While these are natural first metrics, clinical usefulness depends on more than average pointwise error. For glucose forecasting in critical care, performance should also be evaluated in terms of the detection of clinically meaningful excursions, reliability in hypo- and hyperglycemic ranges, calibration of uncertainty, and robustness to missing or noisy sensor measurements. Moreover, because digital twins are intended for integration into decision-support workflows, future assessments should measure computational latency, failure modes, alarm burden, and the extent to which adaptive forecasts improve downstream treatment decisions.

Despite these limitations, the present results suggest a promising path forward for digital twin development in critical care. Rather than relying exclusively on retraining-based adaptation or on static deployment of a pretrained predictor, the proposed workflow illustrates how a foundation model can be paired with a lightweight RL correction layer to achieve continuous, patient-specific updating. This idea is not restricted to glucose forecasting and may be extended to other physiological time series in which a general predictive model provides a strong prior but real-time assimilation is required to track individual trajectories.

## 5 Conclusion

We presented a new digital twin workflow for continuous glucose forecasting that integrates a frozen PatchTST foundation model with an Actor–Critic reinforcement learning agent for bounded residual correction. Unlike our previous transfer-learning-based workflow, which updates the model only after threshold violations, the proposed framework performs proactive, real-time assimilation by adjusting the forecast at each prediction step. This design preserves the global predictive structure of the pretrained foundation model while enabling rapid patient-specific adaptation.

Across both source-data testing and deployment to 10 new ICU patients, the RL-enhanced workflow consistently improved forecasting accuracy over the baseline model. The agent exhibited stable pretraining behavior, reduced the source test MAPE from 0.49% to 0.34%, and lowered the MAPE for every new patient without requiring further retraining. The learned corrections remained within prescribed safety bounds, indicating that improved performance was achieved through controlled and clinically plausible adjustments rather than excessively large corrections.

Taken together, these findings show that residual reinforcement learning provides an effective mechanism for continuous data assimilation in digital twin forecasting. The proposed workflow offers a practical alternative to reactive retraining strategies and provides a flexible foundation for future patient-specific decision-support systems. Future work will focus on larger multi-center validation, incorporation of multimodal clinical inputs, and extension of the RL correction strategy to longer horizons and additional physiological variables.

## Acknowledgments

The authors thank Dr. Hua Jiang’s group for sharing the dataset used in this study. The research of Xiang Cao and Qi Wang is partially supported by NSF awards DMS-2038080 and OIA-2242812; Qi Wang’s research is partially supported by an SC GAIN-CRP award. The funders played no role in study design, data collection, data analysis and interpretation, or manuscript preparation.

## Contributions

X.C. and Q.W. conceived the study. X.C. developed the model, conducted the analyses, validated the model structure and assumptions, and provided input on parameter selection. X.C. and Q.W. interpreted the results and prepared the first draft. Q.W. revised the manuscript and secured funding. All authors contributed to the interpretation of the findings, reviewed the manuscript, and approved the final version.

## Data availability

The datasets used and/or analyzed during the current study are available from the corresponding author on reasonable request.

## Code availability

The code and training and validation datasets used in this study are not publicly available but may be made available to researchers upon reasonable request to the corresponding author.

## References

[1] Xiang Cao, Chao Cai, Jianguo Hou, Xinhang Wei, and Qi Wang. A digital twin for tracking and forecasting glycemia with septic patients in icus. medRxiv, pages 2026–04, 2026.

[2] Xiang Cao, Dong Wang, Jue-Xia He, Jianguo Hou, Jun Li, Hua Jiang, and Qi Wang. Machine learning glucose forecasting models for septic patients. Scientific Reports, 15(1):40432, 2025.

[3] Jorge Corral-Acero, Filippo Margara, Maciej Marciniak, et al. The ‘digital twin’ to enable the vision of precision cardiology. European Heart Journal, 41(48):4556–4564, 2020.

[4] Simon Finfer, Dean R. Chittock, Stephen Y.-S. Su, et al. Intensive versus conventional glucose control in critically ill patients. New England Journal of Medicine, 360(13):1283–1297, 2009.

[5] Simon Finfer, Dean R Chittock, Steve Yu-Shuo Su, Deborah Blair, Denise Foster, Vinay Dhingra, Rinaldo Bellomo, Deborah Cook, Peter Dodek, William R Henderson, Paul C Hébert, Stephane Heritier, Daren K Heyland, Colin McArthur, Ellen McDonald, Imogen Mitchell, John A Myburgh, Robyn Norton, Julie Potter, Bruce G Robinson, and Juan J Ronco. Intensive versus conventional glucose control in critically ill patients. New England Journal of Medicine, 360(13):1283–1297, 2009.

[6] Evangelia Katsoulakis, Qi Wang, Huanmei Wu, Leili Shahriyari, Richard Fletcher, Jinwei Liu, Luke Achenie, Hongfang Liu, Pamela Jackson, Ying Xiao, Tanveer Syeda-Mahmood, Richard Tuli, and Jun Deng. Digital twins for health: a scoping review. npj Digital Medicine, 7:77, 2024.

[7] James S. Krinsley, J. Geoffrey Chase, Jan Gunst, et al. Continuous glucose monitoring in the icu: clinical considerations and consensus. Critical Care, 21(1):197, 2017.

[8] Paul E. Marik and Rinaldo Bellomo. Stress hyperglycemia: an essential survival response! Critical Care, 17(2):305, 2013.

[9] Richard S. Sutton and Andrew G. Barto. Reinforcement Learning: An Introduction. MIT Press, 2 edition, 2018.

[10] Brant H Tudor, Ryan Shargo, Geoffrey M Gray, Jamie L Fierstein, Frederick H Kuo, Robert Burton, Joyce T Johnson, Brandi B Scully, Alfred Asante-Korang, Mohamed A Rehman, and Luis M Ahumada. A scoping review of human digital twins in healthcare applications and usage patterns. npj Digital Medicine, 8:587, 2025.

[11] Chenyang Yu, Jiming Liu, and Shamim Nemati. Reinforcement learning in healthcare: a survey. ACM Computing Surveys, 55(1):1–36, 2021.

